# Clinical meanings of rapid serological assay in patients tested for SARS-Co2 RT-PCR

**DOI:** 10.1101/2020.04.03.20052183

**Authors:** Angelo VirgilioParadiso, Simona De Summa, Daniela Loconsole, Vito Procacci, Anna Sallustio, Francesca Centrone, Nicola Silvestris, Vito Cafagna, Giuseppe De Palma, Antonio Tufaro, Vito Garrisi, Maria Chironna

**Author notes:** correspondence Address for Correspondence Angelo Paradiso, MD, Scientific Direction, IstitutoTumori G Paolo II, IRCCS, Bari, 70125, Italy, Email address. equally contributed (Angelo VirgilioParadiso, Simona De Summa, Vito Garrisi, Maria Chironna).

## Abstract

**Background:** RT-PCR test for identification of viral nucleic acid is the current standard diagnostic method for the diagnosis of COVID-19 disease but technical reasons limit the utilization of this assay onlarge scalescreenings.

**Method:** We verified in a consecutive series of 191 symptomatic patients the clinical information that new rapid serological colorimetric test qualitatively analyzing IgM/IgG expression can provide with respect to standard assay and with respect to clinical outcome of patients.

**Results:** Rapid serological test showed a sensitivity of 30% and a specificity of 89% with respect to the standard assay but, interestingly, these performances improve after 8 days of symptoms appearance. After 10 days of symptoms the predictive value of rapid serological test is higher than that of standardassay. When the behaviour of the two immunoglobulins was evaluated with respect to time length of symptoms appaerance, no significant difference in immunoglobulins behaviour was shown.

**Conclusions:** The rapid serological test analyzed in the present study is candidate to provide information on immunoreaction of the subject to COVID-19 exposure.

## Background

Recently, a novel coronavirus was first reported to cause lethal pneumonia in humans and inter-person transmission in China.[1]. Subsequent molecular studies confirmed that the origin of the transmissible pneumonia was due to a novel coronavirus named SARS-CoV-2 causing the new corona virus disease COVID-19 (2).

As the COVID-19 disease rapidly spread to other Asian and European countries, the Italian Governmenthad to take drastic measures to contain the outbreak. The actions rolled outhave included establishing strict criteria to define patients from whomoropharingeal swabsshould be collected for molecular PCR diagnosis of Covid-19 and to quarantine individualswho may have contacted infected people (3).

These measures have been active since some 4 weeks but the trend of new SARS-CoV-2 infected casesin Italy is still increasing with more than 4000 new casesdaily (4).

Several attempts have been made to interpret the epidemiological trend of Covid-19 in Italy and experts have pinpointed the importance of the limited possibilityof making an early diagnosis of the infection (5) as well as the impossibility to detect asymptomatic subjects carrying the virus (6).

The RT-PCR test for identificationof viral nucleic acid is the current standard diagnostic method for the diagnosis of COVID-19. However, this assay has some practical limitations (3)such as the annoying method to obtain biological material from the nasopharynx, the relatively long time to generate results, the need for certified laboratories and specific expertise. These limitations make RT-PCR unsuitable for a quick and simple patient screening and therefore, the search for a precise, rapid and simple test to quickly identify SARS-CoV-2 infected patients in a large scale screening has become urgent to prevent virus transmission and to ensure timely treatment of patients.

The Saw Swee Hock School of Public Health of the National University of Singapore (https://sph.nus.edu.sg/wp-content/uploads/2020/03/COVID-19-Science-Report-Diagnostics-13-Mar.pdf) recently reviewed the diagnostic test for COVID-19 infection currently undergoing clinical validation by listing dozens of assays based on RT-PcR, RealtimePcR, NGS, Microfluidics. Twelve immunoassays based on the evidence of COVID-19 related IgG andIgMwere also listed. This latter experimental attempt, based on previous experiences with epidemic viral SARS infection, argued that IgM specific SARS-CoV-2 antibody could be detected in the blood after 3-6 days while IgGsome days later (4). It has also been speculated that, since SARS- CoV-2 belongs to the same large family of viruses that caused the MERS and SARS epidemics, their antibody seroconversion should be similar (5).

In the previously mentioned report by Singapore National University, an immunoassay described as with available information regarding sensitivity and specificity is Viva-Diag™ kit produced by Jiangsu Medomics Medical Technologies kit (https://www.vivachek.com/vivachek/English/prods/prod-covid19.html) which, according to the preliminary data available (6), may be a potential candidate for reliable and rapid (15 minutes) testing. The test has been reported to be based on the utilization of Anti-human IgG and anti IgM against the recombinant antigen representing the Receptor-Binding Domain of the COVID-19 spike protein.

Based on these assumptions, we designed a study to verify the clinical information that the serological VivaDiag™ test can provide compared to the standard RT –PCR lab test for SARS-CoV-2 andregarding the clinical outcome of patients.To this purpose, we set upa prospective, mono-institutional, ad hoc, blind and independent study enrolling a series of 191 patients.

## Material and methods

Between March 23^rd^ and March 29^th^, a consecutive cohort of 191 patients having access to Emergency Room of Ospedale Policlinico Consorziale of Bari for Covid-19 disease orienting-symptoms has been enrolled. All these patients received oropharingeal swap for standard SARS-CoV-2 rt-PCR analysis and, simultaneously venous blood sampling for Viva-Diag™ test performance. Registries and main clinical information comprising timing of appearance of symptoms were collected togheter with Informed Consent. Oropharingeal swap samples were suddenly analyzed for SARS-CoV-2 rt-PCR in Laboratory of Molecular Epidemiology and Public Health of the Hygiene Unit of Policlinico Hospital (Bari, Italy), which is the Regional Reference Laboratory for the SARS-CoV-2 identification. Venous blood sampled were analyzed in Clinical Pathology Laboratory (Certified ISO- 9001/2015;Head E. Savino) and Institutional Biobank (Certified ISO- 9001/2015;Head A. Paradiso) of Istituto Tumori G Paolo II, IRCCS, Bari (I). The study has been approved by Ethical Committee of IstitutoTumori G Paolo II, IRCCS, Bari with Protocol number CE 870/2020.

### Molecular detection of SARS-CoV-2

Nasopharyngeal/oropharyngeal swabs weresubjected to nucleic acid extraction by MagNA Pure (Roche Diagnostics, Mannheim, Germany), according to the manufacturer’s instructions. The presence of E gene, RdRP gene and N gene of SARS-CoV-2 virus were identified by a commercial real-time PCR assay (Allplex2019-nCoV Assay; Seegene, Seoul, Republic of Korea). Samples were considered positive at molecular screening if all the three genes were detected. Moreover, the WHO Real-time rRT-PCR protocol was used to confirm the presence of SARS-CoV-2 (https://www.who.int/docs/default-source/coronaviruse/uscdcrt-pcr-panel-for-detection-instructions.pdf?sfvrsn=3aa07934_2).

### SARS-CoV-2 rapid IgG-IgM Test

SARS-CoV-2 rapid IgG-IgM Viva-Diag™Test combined antibody test kit designed and manufactured by Jiangsu Medomics Medical Technologies.The test is based on a lateral flow qualitative immunoassay for the rapid determination of the presence or absence of both anti- SARS-CoV-2-IgM and anti- SARS-CoV-2-IgG in human specimens (whole blood, serum, and plasma). A surface antigen from SARS-CoV-2 which can specifically bind to SARS-CoV-2 antibodies (including both IgM and IgG) is conjugated to colloidal gold nanoparticles and sprayed on conjugation pads. The SARS-CoV-2 rapid IgG-IgM combined antibody test strip has two mouse anti-human monoclonal antibodies (anti-IgG and anti-IgM) stripped on two separated test lines.

When testing, 10-15ul of specimen was added into the sample port followed by the addition of sample dilution buffer. As the specimen flows through the device, anti-SARS-CoV-2 IgG and IgM antibodies, if present in the specimen, are bound by the SARS-CoV-2 antigen labeled gold colorimetric reagent fixed on the conjugate pad. As the conjugated sample continues to travel up the strip, the anti- SARS-CoV-2 IgM antibodies are bound on the M(IgM) line, and the anti-COVID-19 IgG antibodies are bound to the G (IgG) line. If the specimen does not contain SARS-CoV-2 antibodies, no labeled complexes bind at the Test. The presence of SARS-CoV-2 IgG and IgM antibodies are indicated by a red/purple line in the specific region indicated on the device.Each test was evaluated by two operators and picture was taken. In case of disagreement, the picture was evaluated by a third party.

### Statistical analyses

The performance of theVivaDiag™tests was compared to that of the RT-PCR tests via the “caret” R package that computed all the parameters needed (accuracy, precision, recall, kappa). Univariate and multivariate logistic regression was performed. Age was dichotomized using the median as a cut-off; days from the onset of symptoms was also transformed into a categorical variable (0-5 days, 6-8 days, 9- 10 days, 11-15 days, more than 15 days). All the analyses were carried outin R v3.6 and results were considered to be significant when the p-value was <0.05.

## Results

All the 191 subjects enrolled in the study had a SARS-CoV-2 RT-PCR test and RapidIgG/IgM test performed. The cohort had a median age of 58.5 years, 60.62% were male and had presented to the Emergency Room at different times from the onset of symptoms. A description of the symptoms wasavailable in 160 of the191 patients. Fourteen subjects (8.7%) were asymptomatic at time the came to the Emergency Room.

70 patients (37%) had positive SARS-CoV-2 RT-PCR tests while 33 (18%) had positive serum IgM/IgGtests. Compared to the RT-PCRtest, the serological test showed an accuracy of 0.67 (95%CI: 0.604-0.741), a sensitivity of 30% and a specificity of 0.89%. Cohen’s Kappa value was 0.21, whose strength of agreement, according to Altman (10) was considered “Fair”. Notably,13 patients (7%) had positiveserological testsbut negative RT-PCRresult.

The distribution of the percentage of positive results detected by both tests broken down by days from symptom onset is shown in Figure 1. Aclear increase inpositive serological tests can be observed as more days elapsed from symptom appearance, reaching 66.67% 15 days after symptom onset. Conversely, the highest likelihood of a positive RT-PCR test resultwas seen9-10 days from symptom onset and it decreased after that point in time. Amongstthe asymptomatic individuals,28.57% had positive RT-PCR test result while21.43% had serological test one.

**Figure 1.**
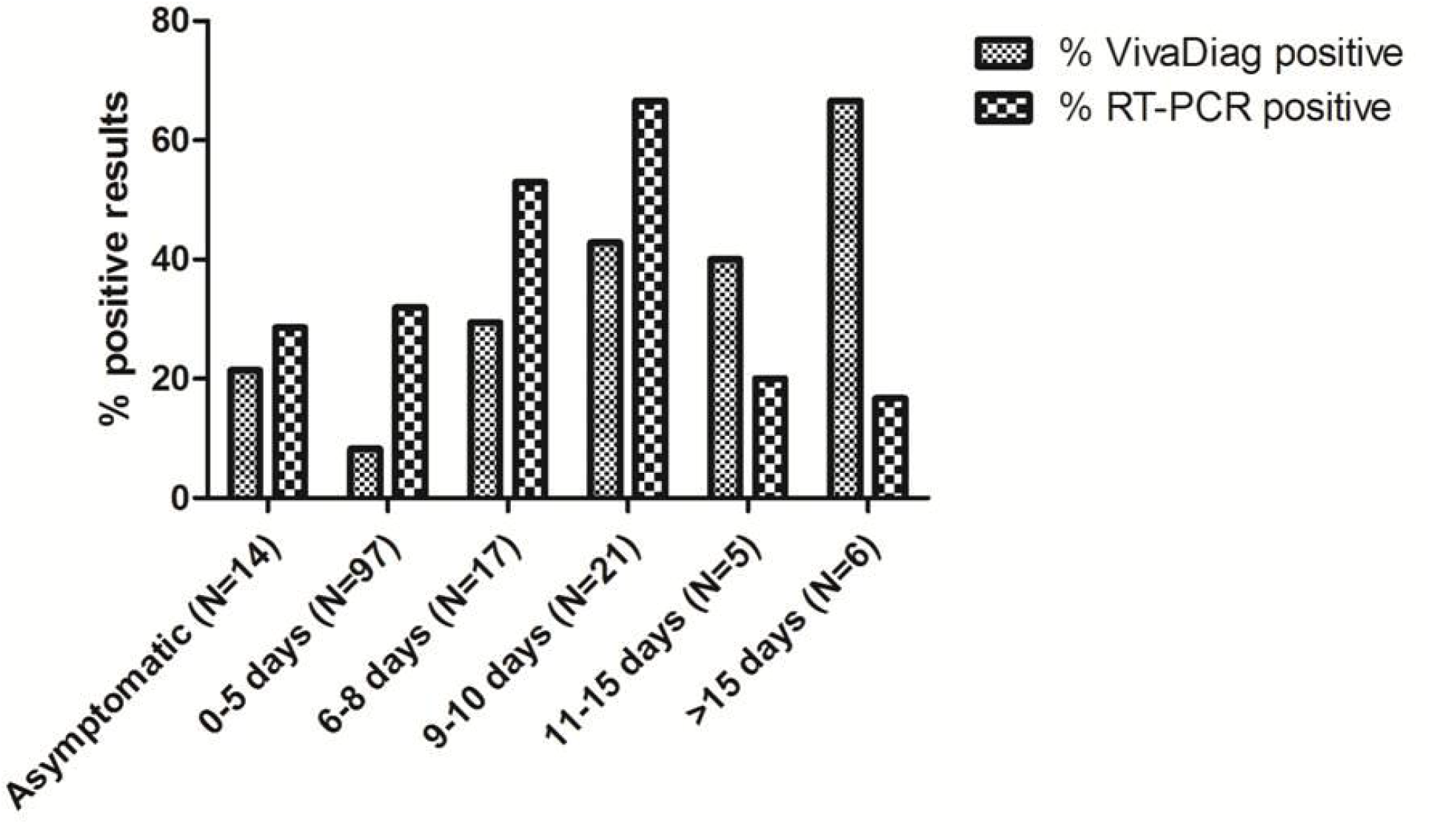
Barplot representing the distribution of proportion of positive results along time-points from symptom onset (including asymptomatic group)

A further analysis regardingthe behavior of IgM and IgGaccording to the time from symptom onset is described in Figure 2. Only minimal differences in the behavior of the two immunoglobulins with respect tothe time of symptom appearance became evident. However, all 13 patients withVivaDiag™ positive test and negative RT-PCR result (6.8%) were positive for IgM while only 7 subjects resulted positive for IgG.

**Figure 2.**
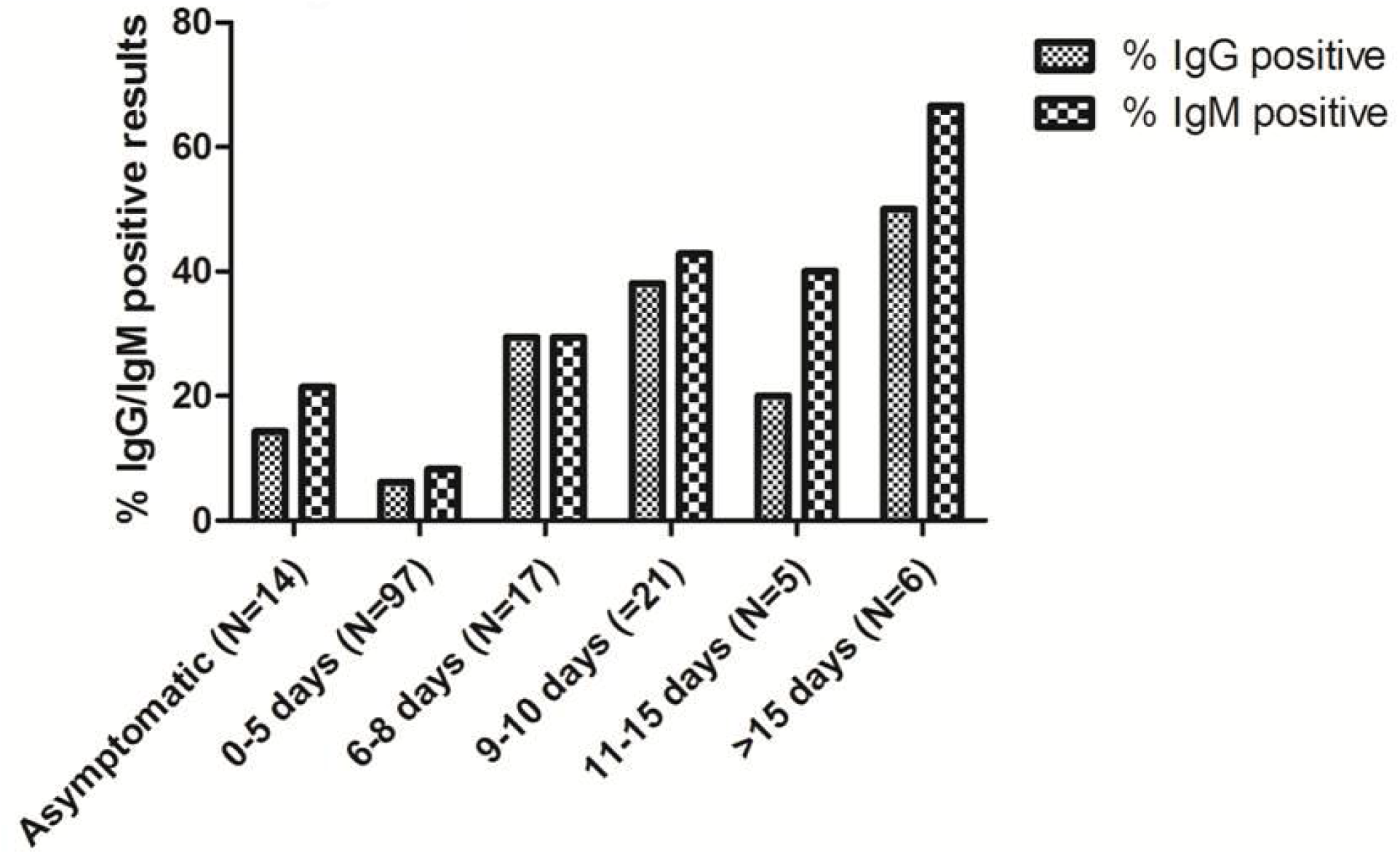
Barplot representing the distribution of the percentage of positive results distinguishing IgG and IgM results from VivaDiag™test.

Univariate and multivariate logistic regressions were performedto look for independent variables predictive of positive VivaDiag™ and RT-PCR test results (Table 2).Both univariate and multivariate analysesshowed that age>58.5years and more than 15 days from symptom onset were significantly associated with VivaDiag™positivity while 9-10 days after symptom onset was related to positive RT-PCR test result.

**TABLE 1.**

|  |  | **RT-PCR** |  | *p-value* |
| --- | --- | --- | --- | --- |
|  |  | **NEGATIVE** | **POSITIVE** |  |
| **VivaDiag™** | **NEGATIVE** | 107 (56.31%) | 49 (28.78%) | 0.001 |
|  | **POSITIVE** | 13 (6.84%) | 21 (11.05%) |  |

**TABLE 2.**

| **VivaDiag™ uni variate logistic regression** |  |  |
| --- | --- | --- |
|  | OR (95%CI) | P-value |
| **Days from symptoms** |  |  |
| Asymptomatic | Ref | Ref |
| 0-5 | 0.32 (0.08-1.66) | 0.13 |
| 6-8 | 1.52 (0.3-8.9) | 0.61 |
| 9-10 | 2.74 (0.63-14.88) | 0.19 |
| 11-15 | 2.44 (0.23-23.39) | 0.42 |
| >15 | 7.33 (0.96-77.28) | 0.06 |
| **Age** |  |  |
| ≤58.5 | Ref | Ref |
| >58.5 | 2.99 (1.31-7.31) | 0.01 |
| **Sex** |  |  |
| F | Ref | Ref |
| M | 1.22 (0.55-2.85) | 0.62 |
| **VivaDiag™ multivariate logistic regression** |  |  |
| **Age** |  |  |
| ≤58.5 | Ref | Ref |
| >58.5 | 3.59 (1.39-10.48) | 0.01 |
| **Days from symptoms** |  |  |
| >15 | 12.3 (1.44-148.14) | 0.02 |
| **RT-PCR uni variate logistic regression** |  |  |
| **Days from symptoms** |  |  |
| Asymptomatic | Ref | Ref |
| 0-5 | 1.17 (0.36-4.54) | 0.79 |
| 6-8 | 2.81(0.65-13.75) | 0.17 |
| 9-10 | 4.99 (1.21-24.11) | 0.03 |
| 11-15 | 0.62 (0.02-6.17) | 0.71 |
| >15 | 0.53 (0.02-4.64) | 0.57 |
| **Age** |  |  |
| ≤58.5 | Ref |  |
| >58.5 | 0.89 (0.47-1.7) | 0.74 |
| **Sex** |  |  |
| F | Ref | Ref |
| M | 1.2 (0.62-2.33) | 0.58 |
| **RT-PCR multivariate logistic regression** |  |  |
| **Days from symptoms** |  |  |
| 9-10 | 4.96 (1.2-24) | 0.03 |

## Discussion

The clinical relevance of so-called rapid serological testing is still an open issue since the data currently available are still scarce (9). For this reason we have analyzed its performance compared to standard RT-PCR testing and with respect to the time of onset of Covid-19 related symptoms. To this end, we set up a mono- institutional consecutive cohort of patients who were tested with both assays each in a single qualified laboratory.

The design of our study allowed us to specifically analyze two aspects of the open issue: the concordance of the rapid serological test with standard molecular testing; the trend of immunoglobulinsIgG/IgMexpression with respect to the onset of clinical symptoms.

Regarding the degree of concordance between the two tests, the results reported in Table 1 clearly show that the precision of the rapidserological test Viva-Diag™is unsatisfactory. Notably, only 43% of the patients that were COVID-19-positive with the molecular test were also positive with serological test. This figure is impressively similar to the performances reported for serological tests in Spain (11) and Germany (12). However, the first important information from our study concerns the 12% of cases that tested negative with molecular test but had positive serological results. The two tests do not produce similar results as would be obvious for assays that look at different aspects of Covid-19; in fact, the molecular test demonstrates the presence of SARS-CoV-2 virus in samples from specific anatomical parts of the respiratory system while the kinetics of immunoglobulinsis devoted to describe as the body reacts to the viral infection. Negative serological test results in patients with a positive molecular test could mean that the latter are infected subjects that have not yet reached the stage of developing an immunoglobulin reaction; conversely, subjects whose serological tests show the presence of specific IgG and/or IgM antibodies and have negative molecular tests, may be considered as recovering from COVID-19. The data shown in Figure 1 seem to confirm these assumptions as the molecular test yielded more positive results in early symptomatic phases of the disease while the serological test performed better in the late phases of the disease (i.e. 10 days after symptom appearance).

The second aspect we were able to analyze in our study regarded seroconversion and the kinetics of immunoglobulins with respect to the onset of Covid-19 related symptoms. Figure 2 shows the behavior of the two immunoglobulins with respect to symptom appearance. Interestingly, IgG and IgM do not seem to behave differently based on the number of days elapsing from symptom appearance but they display a clear progressive increase along the course of the disease. This unexpected finding, in contrast with common thinking concerning the kinetics of the two immunoglobulins, is supported by the results reported by Zhang (13) and Lou (14) reportingthat detectable serology markers IgG and IgM had a similar seroconversion in COVID-19 patients with antibody levels increasing rapidly starting from 6 days post exposure and that this trend occurred with a concomitant decline in viral load. Such a behavior in the 6-10 day time window after symptom appearance is accompanied by an improvement in serological test sensitivity compared to standard molecular testing

Our study has some important limitations. Firstly, the Viva-Diag™ test is based on a colorimetric evaluation of the IgG and IgM bands by the operator thus implying all the limitations that a qualitative inter-intra-operator evaluation produce in terms of variability (15). In our study, this aspect was partially solved by resorting to double operator evaluation and taking picture of all test results to be re-analyzed by a third party in the case of first level evaluation disagreement. However, in this regard, our next step of research to overcome the issues met,will be to use quantitative immunoenzymatic methods to analyze SARS-CoV-2 specific immunoglobulins (16).

A further limitation of our study regarded the fact that the neutralizing antibodies used in the Viva-diag™ test might cross-react with other corona-virus antigens, like those of the SARS-CoV. The recombinant antigen utilized in Viva-Diagis the receptor binding domain of SARS-CoV-2 spike protein for which information on possible cross-reactivity with other corona and flu viruses has not yet been studied (9). Further studies are urgently needed to definitely clarify this point.

## Conclusions

Our study analyzed theclinical performance of the rapid serological test, Viva- Diag™ and confirmedthe test’s limited applicability for the diagnosis of SARS-CoV- 2 infection when compared to standard molecular testing. However, this rapid serological test seems to provide important information concerning the immunoreaction of individuals to the infection and, more importantly, it may detect previous exposure to the virus in currently healthy persons. The trial, recently registered in ClinicalTrial.gov (NCT04316728), specifically addresses this issue by planning to investigate the monitoringof seroconversion of COVID-19 IgG/IgM in healthy subjects who may develop COVID-19 related symptoms.

## Data Availability

All relevant data are displayed in the manuscript.

## Acknowledgments

The AA wish to thank Clinical pathology Laboratory Director, Dr E Savino and Coworkers for the cohoperation.

## Reference list

1) Huang C, Wang Y, Li X, Ren L, Zhao J, Hu Y, Zhang L, Fan G, Xu J, Gu X,Cheng Z, Yu T, Xia J, Wei Y, Wu W, Xie X, Yin W, Li H, Liu M, Xiao Y, Gao H, Guo L,Xie J, Wang G,Jiang R, Gao Z, Jin Q, Wang J, Cao B. Clinicalfeatures of patientsinfected with 2019 novel coronavirus in Wuhan, China. Lancet 2020: 395(10223): 497–06

2) Zhu N, Zhang D, Wang W, Li X, Yang B, Song J, Zhao X, Huang B, Shi W, Lu R, Niu P, Zhan F, Ma X, Wang D, Xu W, Wu G, Gao GF, Tan W; China Novel Coronavirus Investigating and Research Team. A Novel Coronavirus from Patients with Pneumonia in China, 2019. N Engl J Med. 2020 Feb 20;382(8):727–733. doi: 10.1056/NEJMoa2001017. Epub 2020 Jan 24. PubMed PMID: 31978945.

3) Livingston E, Bucher K. Coronavirus Disease 2019 (COVID-19) in Italy. JAMA. 2020 Mar 17. doi: 10.1001/jama.2020.4344. [Epub ahead of print] PubMed PMID: 32181795.

4) https://www.worldometers.info/coronavirus/country/italy/ (April, 1, 2020)

5) Jin YH, Cai L, Cheng ZS, et al. A rapid advice guideline for the diagnosis and treatment of 2019 novelcoronavirus (COVID-19) infectedpneumonia (standard version). Mil Med Res. 2020; 7 (1): 4

6) Rothe C, Schunk M, Sothmann P et al, Transmission of COVID-19 infection from an asymptomaticcontact in Germany. N Engl. J Medicine, 2020

7) Lee HK, Lee BH, Seok SH, et al. Production of specific antibodies against SARS-Coronavirus nucleocapsid protein without cross reactivity with human coronaviruses 229E and OC43. J Vet Sci. 2010; 11(2): 165–167

8) Tian X, Li C, Huang A, Xia S, Lu S, Shi Z, Lu L, Jiang S, Yang Z, Wu Y, Ying T. Potent binding of 2019 novel coronavirus spike protein by a SARS coronavirus-specific human monoclonal antibody. Emerg Microbes Infect. 2020 Dec;9(1):382–38

9) Li Z, Yi Y, Luo X, Xiong N, Liu Y, Li S, Sun R, Wang Y, Hu B, Chen W, Zhang Y, Wang J, Huang B, Lin Y, Yang J, Cai W, Wang X, Cheng J, Chen Z, Sun K, Pan W, Zhan Z, Chen L, Ye F. Development and Clinical Application of A Rapid IgM-IgG Combined Antibody Test for SARS-CoV-2 Infection Diagnosis. J Med Virol. 2020 Feb 27

10) Landis, J.R.; Koch, G.G. (1977). “The measurement of observer agreement for categorical data”. Biometrics. 33 (1): 159–174

11) https://english.elpais.com/society/2020-03-27/unreliability-of-new-tests-delays-effort-to-slow-coronavirus-spread-in-spain.html

12) https://www.faz.net/aktuell/gesellschaft/gesundheit/coronavirus/neue-corona-symptome-entdeckt-virologe-hendrik-streeck-zum-virus-16681450.html

13) Jin Zhang, Jianhua Liu, Na Li, Yong Liu, Rui Ye, Xiaosong Qin, Rui Zheng. Serological detection of 2019-nCoV respondto the epidemic: A useful complement to nucleic acid testing https://doi.org/10.1101/2020.03.04.20030916

14) Bin Lou, Ting-Dong Li, Shu-Fa Zheng, Ying-Ying Su, Zhi-Yong Li, Wei Liu, Fei Yu, Sheng-Xiang Ge, Qian-Da Zou, Quan Yuan, Sha Lin, Cong-Ming Hong, Xiang-Yang Yao, Xue-Jie Zhang, Ding-Hui Wu, Guo-Liang Zhou, Wang-Heng Hou, Ting-Ting Li, Ya-Li Zhang, Shi-Yin Zhang, Jian Fan, Jun Zhang, Ning-Shao Xia and Yu Chen. Serology characteristics of SARS-CoV-2 infection since the exposure and post symptoms onset https://doi.org/10.1101/2020.03.23.20041707

15) Paradiso A, Ellis IO, Zito FA, Marubini E, Pizzamiglio S, Verderio P. Short-and long-term effects of a training session on pathologists’ performance: the INQAT experience for histological grading in breast cancer. J Clin Pathol. 2009 Mar;62(3):279–81. doi: 10.1136/jcp.2008.061036. PubMed PMID: 19251956

16) Guo L, Ren L, Yang S, Xiao M, Chang, Yang F, Dela Cruz CS, Wang Y, Wu C, Xiao Y, Zhang L, Han L, Dang S, Xu Y, Yang Q, Xu S, Zhu H, Xu Y, Jin Q, Sharma L, Wang L, Wang J. ProfilingEarlyHumoralResponse to DiagnoseNovel Coronavirus Disease(COVID-19). Clin Infect Dis. 2020 Mar 21. pii: ciaa310. doi: 10.1093/cid/ciaa310. [Epub ahead of print] PubMed PMID: 32198501

